# Volumetric Analysis of Acute Uncomplicated Type B Aortic Dissection Using an Automated Deep Learning Aortic Zone Segmentation Model

**DOI:** 10.1101/2024.03.29.24305035

**Authors:** Jonathan R. Krebs, Muhammad Imran, Brian Fazzone, Chelsea Viscardi, Benjamin Berwick, Griffin Stinson, Evans Heithaus, Gilbert R. Upchurch, Wei Shao, Michol A. Cooper

**Affiliations:** Department of Surgery, Division of Vascular Surgery, University of Florida, Gainesville, FL; Department of Medicine, University of Florida, Gainesville, FL; Department of Radiology, University of Florida, Gainesville, FL

## Abstract

**Introduction:** Machine learning techniques have shown excellent performance in 3D medical image analysis, but have not been applied to acute uncomplicated type B aortic dissection (auTBAD) utilizing SVS/STS-defined aortic zones. The purpose of this study was to establish a trained, automatic machine learning aortic zone segmentation model to facilitate performance of an aortic zone volumetric comparison between auTBAD patients based on rate of aortic growth.

**Methods:** Patients with auTBAD and serial imaging were identified. For each patient, imaging characteristics from two CT scans were analyzed: (1) the baseline CTA at index admission, and (2) either the most recent surveillance CTA, or the most recent CTA prior to an aortic intervention. Patients were stratified into two comparative groups based on aortic growth: rapid growth (diameter increase ≥5mm/year) and no/slow growth (diameter increase <5mm/year).

Deidentified images were imported into an open-source software package for medical image analysis and randomly partitioned into training(80%), validation(10%), and testing(10%) cohorts. Training datasets were manually segmented based on SVS/STS criteria. A custom segmentation framework was used to generate the predicted segmentation output and aortic zone volumes.

**Results:** Of 59 patients identified for inclusion, rapid growth was observed in 33 (56%) patients and no/slow growth was observed in 26 (44%) patients. There were no differences in baseline demographics, comorbidities, admission mean arterial pressure, number of discharge antihypertensives, or high-risk imaging characteristics between groups (p>0.05 for all). Median duration between baseline and interval CT was 1.07 years (IQR 0.38-2.57). Post-discharge aortic intervention was performed in 13 (22%) of patients at a mean of 1.5±1.2 years, with no difference between groups (p>0.05). In both groups, all zones of the thoracic and abdominal aorta increased in volume over time, with the largest relative increase in Zone 5 with a median 24% increase (IQR 4.4-37). Baseline zone 3 volumes were larger in the no/slow growth (6v^3^) than the rapid growth group (5v^3^) (p=0.03). There were no other differences in baseline zone volumes between groups (p>0.05 for all). Dice coefficient, a performance measure of the model output, was 0.73. Performance was best in Zones 4 (0.82), 5(0.88), and 9(0.91).

**Conclusions:** To our knowledge this is the first description of an automatic deep learning segmentation model incorporating SVS-defined aortic zones. The open-source, trained model demonstrates high concordance to the manually segmented aortas with the strongest performance in zones 4, 5, and 9, providing a framework for further clinical applications. In our limited sample, there were no differences in baseline aortic zone volumes between rapid growth and no/slow growth patients.

**ARTICLE HIGHLIGHTS:** *Type of Research:* Single-center retrospective cohort study

*Key Findings:* A deep learning model was developed to analyze volumetric growth in patients with medically managed acute uncomplicated type B aortic dissection. Volumetric growth was most pronounced in zones 5 (24%), 4 (13%), and 3 (11%). Model performance was best in zones 4, 5, and 9.

*Take Home Message:* A trained, automated, open-source aortic zone segmentation model can accurately track changes in aortic growth by zone over time, providing framework for further clinical applications.

*Table of Contents Summary:* A trained, automated model was developed to analyze aortic zone volumetric growth in a retrospective study of 59 patients with medically managed acute uncomplicated TBAD. Volumetric growth was most pronounced in zones 3-5, while model performance was best in zones 4,5, and 9.

## Background

Despite 20 years of international clinical trials and evaluation, there is no clear data to guide the optimal treatment for patients presenting with acute uncomplicated type B aortic dissection (auTBAD). Traditional management with anti-hypertensive therapy and imaging surveillance results in acceptable early outcomes but long-term survival is poor and aortic degeneration mandating intervention occurs in 40-60% of patients.^1,2^ Thoracic endovascular aortic repair (TEVAR) in the subacute period has emerged as a viable alternative that may improve aortic remodeling and decrease TBAD-related mortality, but carries an increased risk of procedural complications, higher cost, and risk of overtreatment.^3,4^

Due to the clinical equipoise between these two treatment modalities, a qualitative assessment of dissection morphology often drives the initial decision to perform TEVAR in the acute/subacute period. While several high-risk radiographic parameters have been suggested including false lumen diameter >22mm, presence of a lesser curve entry tear, and radiographic malperfusion, aortic diameter >40mm is the only prospectively validated high-risk feature predicting need for future intervention.^5–9^ Additionally, although diameter measurements are the gold standard used to evaluate aortic growth over time, they are subject to high inter-reader variability and do not capture the three-dimensional (3D) nature of aortic growth.^10^ Volumetric CT angiography (CTA) has been suggested as a means of overcoming the limitations of diameter-based measurements and may be better suited to track 3D aortic growth over time.^11,12^ Deep learning techniques have been applied to 3D medical image analysis, with particularly strong performance demonstrated in the computer-aided detection of abnormal mammograms and the identification of lung nodules, for example.^13–16^ In aortic disease, deep learning techniques have been used to model aortic dissections, but none have used the output to examine changes in total aortic growth over time.^17^ In addition, no prior studies have examined volumetric growth patterns across aortic zones defined by Society for Vascular Surgery (SVS) and Society of Thoracic Surgeons (STS) reporting guidelines, which may demonstrate distinct behavior after dissection (**Figure 1**).^9^The purpose of this study was to establish a trained, automated deep learning aortic zone segmentation model to facilitate performance of a volumetric analysis of patients with medically managed auTBAD. A secondary objective was to determine if differences in baseline aortic zone volumes were associated with aortic growth rate over time. We hypothesized that baseline thoracic aortic volumes would be higher in patients that experienced aortic growth over time compared to those that did not.

**Figure 1:**
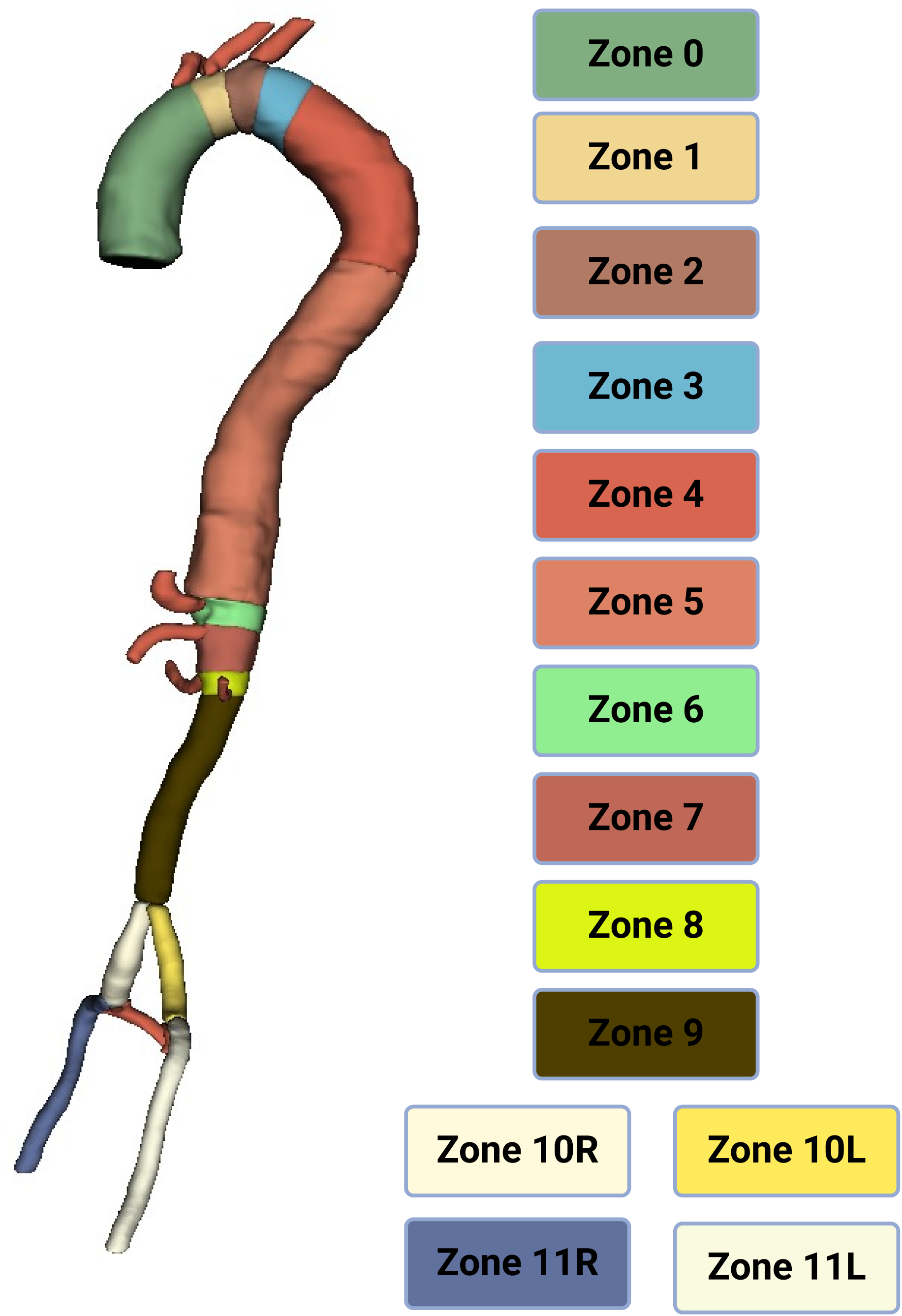
SVS/STS defined aortic zones in relation to primary aortic branches. Zone 0: aortic root to innominate artery. Zone 1: innominate artery to left common carotid artery. Zone 2: left common carotid to left subclavian artery. Zone 3: first 2cm distal to the left subclavian artery. Zone 4: Zone 3 to T6 vertebral body. Zone 5: Zone 4 to celiac artery. Zone 6: celiac artery to superior mesenteric artery. Zone 7: superior mesenteric artery to most proximal renal artery. Zone 8: renal to infra-renal abdominal aorta. Zone 9: Infrarenal abdominal aorta. Zone 10: common iliac arteries. Zone 11: external iliac arteries.

## Methods

### Patient identification

The study was approved with a waiver of consent from our Institutional Review Board (IRB 202100962). Using a prospectively maintained institutional database, a retrospective review of patients admitted to our center with a diagnosis of code of aortic dissection (ICD-9 codes 441.01, 441.03; ICD-10 codes I71.00, I71.01) between 10/2011 and 3/2020 was performed. Type A dissection, intramural hematoma, penetrating aortic ulcer, and chronic aortic dissection patients were excluded. TBAD patients that underwent TEVAR on their index admission were excluded. Uncomplicated TBAD patients were identified based on the absence of malperfusion, rupture, rapid degeneration, or refractory pain. All uncomplicated TBAD patients were medically managed without surgical intervention at index admission.

Imaging was then reviewed and patients without high-resolution surveillance imaging beyond three months from discharge after index hospitalization were excluded. Patients were reviewed to ensure high-resolution surveillance CTA (≤3mm slices) at both diagnosis and surveillance. Given that many initial CTAs were sourced from various local imaging centers with differing protocols, our study’s criteria mandated that images were captured in ≤3mm slices. Mean number of axial slices was 734, with mean slice thickness 0.969 mm. To optimize our 3D training model, patients with bovine arch or aberrant arch anatomy were excluded.

### Data collection

Patient demographics, comorbidities, and hospital course were obtained from the electronic medical record. A radiologist and vascular surgeon analyzed imaging characteristics from two CT scans: the baseline CTA at index admission, and either the most recent surveillance CTA, or the most recent CTA prior to an aortic intervention if one was performed. Imaging characteristics including total aortic diameter, true and false lumen diameter, and high-risk features including presence of lesser curve entry tear and thrombosis status of the false lumen were all assessed. Patients were then stratified into two groups based on maximum aortic diameter changes over time: rapid growth (diameter increase ≥5mm/year and no/slow growth (diameter increase <5mm/year).

### Model Implementation

CT scans were randomly partitioned into training (80%), validation (10%), and testing (10%) cohorts for unbiased evaluation. Our training pipeline was implemented using the PyTorch framework and MONAI library.^18^ To expedite model training, we re-sampled volumes to a uniform spacing and employed a random cropping center to re-sample random patches for training, enhancing data diversity and mitigating overfitting. Our model was trained for 3000 iterations, roughly equivalent to 666 epochs. Training was performed on a single NVIDIA A100 GPU with 80 GB RAM.

### Manual Segmentation

Baseline and surveillance CTAs were deidentified and exported from our institutional software platform and uploaded to 3D Slicer (https://www.slicer.org), a free open-source software package for medical image analysis.^19^ After importing the deidentified images into 3D Slicer, the 11 aortic zones were manually segmented based on SVS/STS criteria. A Gaussian smoothing filter was applied to reduce jaggedness and enhance 3D continuity.

### Segmentation Model

A custom Context Infused Swin-UNet (CIS-UNet) segmentation framework was used for multi-class 3D aortic segmentation (**Figure 2**).^20^ CIS-UNet integrates the capabilities of convolutional neural networks (CNNs) and the Swin transformer to effectively capture both local and global features. CIS-Unet consists of a CNN encoder block architecture, a Swin transformer in the bottleneck layer, and a decoder using transposed CNNs.^20^ CIS-Unet features a novel self-attention block to efficiently capture long range dependencies between image patches. In aortic branch segmentation, CIS-Unet has been shown to outperform state-of-the-art segmentation models.^20^ Using the 3D output generated, the volume in voxels cubed of each aortic zone was computed using the segment statistics function within 3D Slicer.

**Figure 2:**
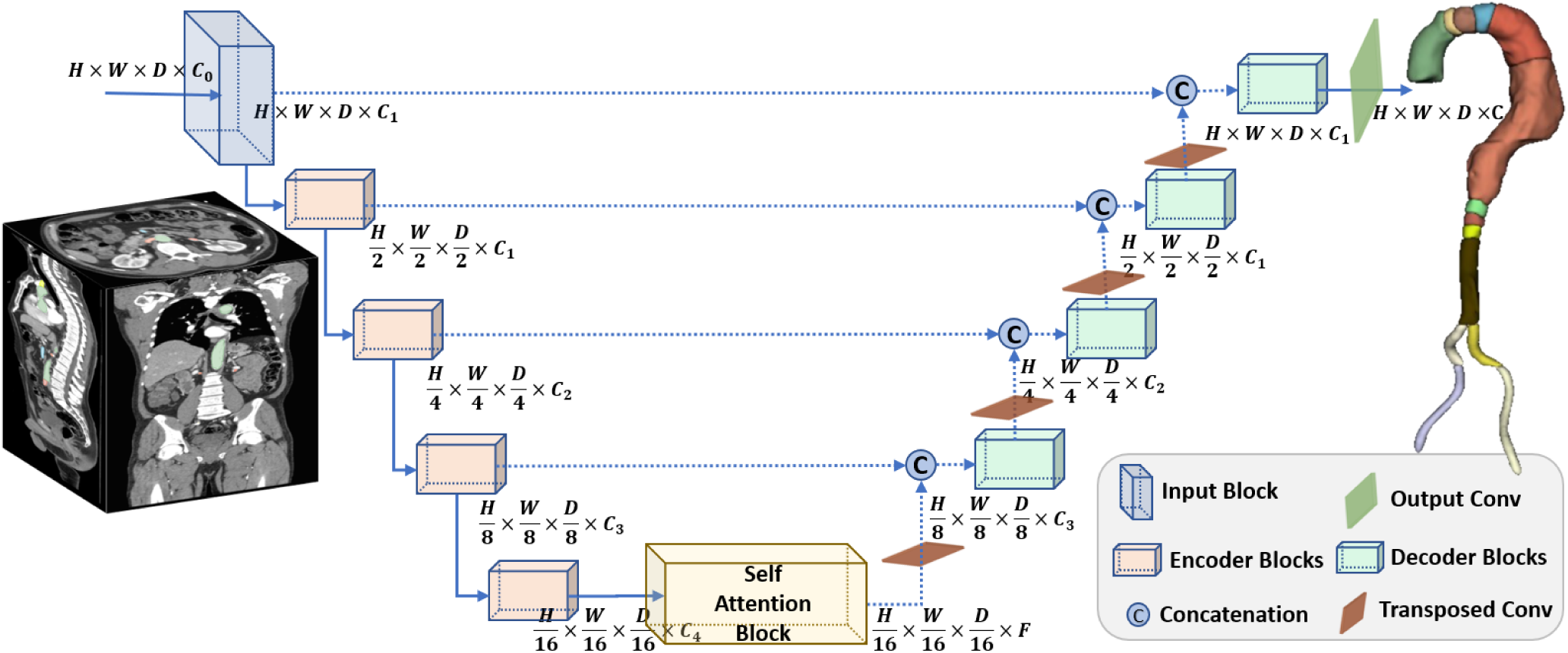
CIS-Unet: a hybrid model used for multi-class segmentation of the aorta that combines convolutional neural networks and vision transformers to capture both local and global image features to enhance segmentation accuracy.

### Statistical Analysis

Model performance was assessed by mean Dice coefficient. Primary comparisons were made of the volumes in the different aortic zones between patients with rapid and no/slow aortic growth using R statistical package. Group comparisons were made using t-test/Man-Whitney test and Chi-square/Fisher Exact test as appropriate with a p-value of ≤0.05 considered significant. In addition, percentage increase in zone and whole aorta volume per year ((final volume-initial volume)/(initial volume)*100/surveillance duration) was plotted by patient using Prism 10 software (GraphPad, La Jolla, CA, USA).

## Results

Of the 159 patients treated for uncomplicated TBAD, 76 patients had the requisite surveillance imaging for inclusion. An additional 17 patients were excluded due to aberrant aortic arch anatomy (**Figure 3**). Patient characteristics are shown in **Table I**. Of 59 included patients, average age was 59.2 years and 66% were male. Initial mean arterial pressure (MAP) was 95.5 mmHg. Hypertension was a listed diagnosis in 81% of patients, followed by any tobacco use history (68%) and renal disease (14%). Patients were discharged with an average of 3 antihypertensive medications. Rapid aortic growth was observed in 33 (56%) patients, with no/slow aortic growth observed in 26 (44%) patients. There were no differences in baseline demographics, comorbidities, admission mean arterial pressure, or number of discharge antihypertensives between groups (p>0.05 for all). Median duration between baseline and interval CT was 1.07 years (IQR 0.38-2.57); it was longer in those with no/slow aortic growth (median 2.2 years (IQR 1.2-4.0)) compared to those with rapid aortic growth (median 0.65 years (IQR 0.26-1.09)) (p<0.01). Among all patients, thoracic aortic diameter increased at a median of 4.9 (IQR 1.6-13.1) mm/year. In patients with rapid aortic growth, median annual growth was 11.0 (IQR 5.8-20.9) mm/year, compared to an annual growth rate of 1.7 (IQR 0.1-3.8) mm/year in patients with no/slow aortic growth (p<0.01). Post-discharge aortic intervention was performed in 13 (22%) of patients during the surveillance period at a mean of 1.5±1.2 years, with no difference in incidence between patients with rapid aortic growth (n=7, 21%) and no/slow aortic growth (n=6, 23%) (p=1).

**Figure 3:**
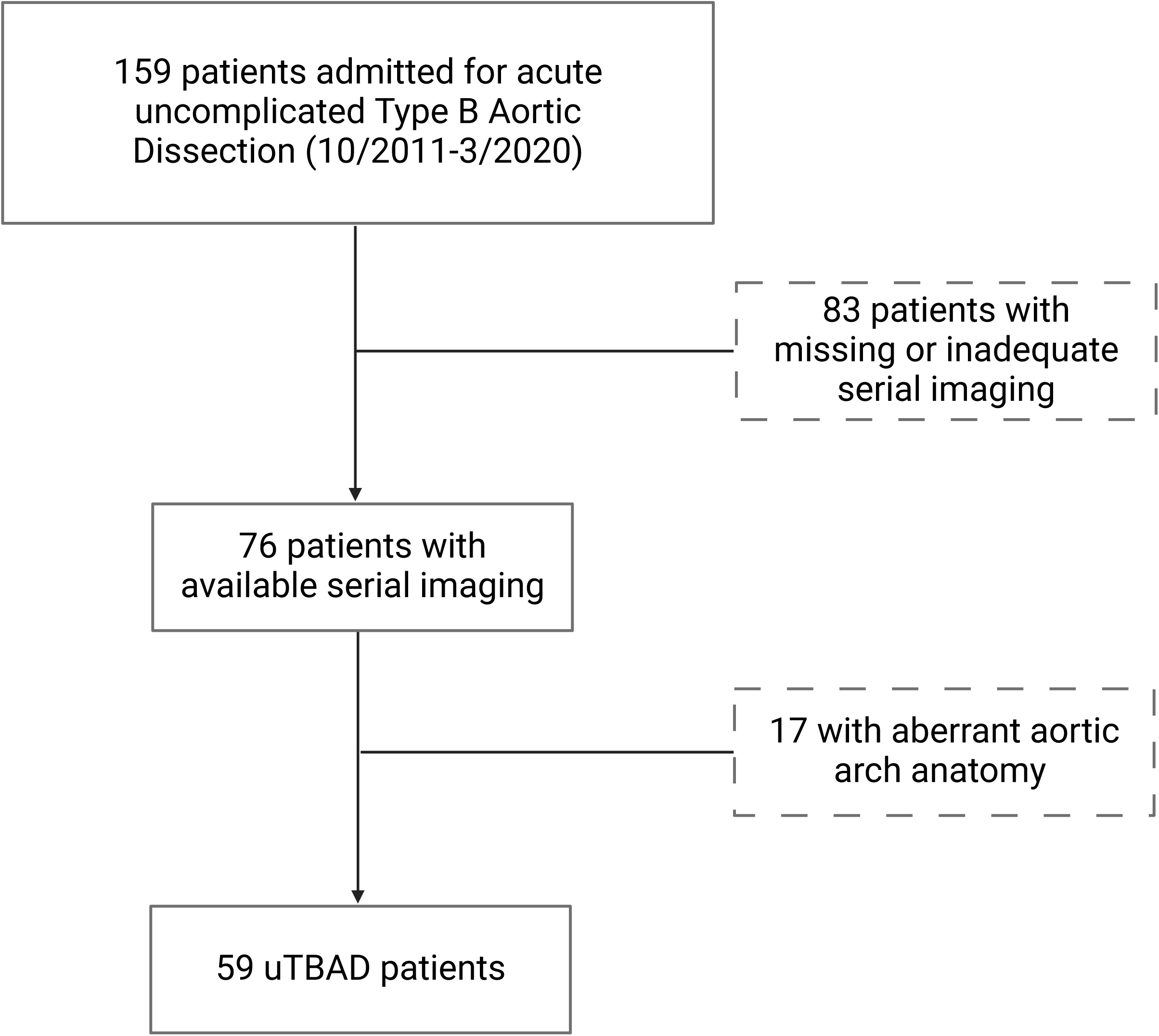
Patient selection flowchart.

**Table I:**
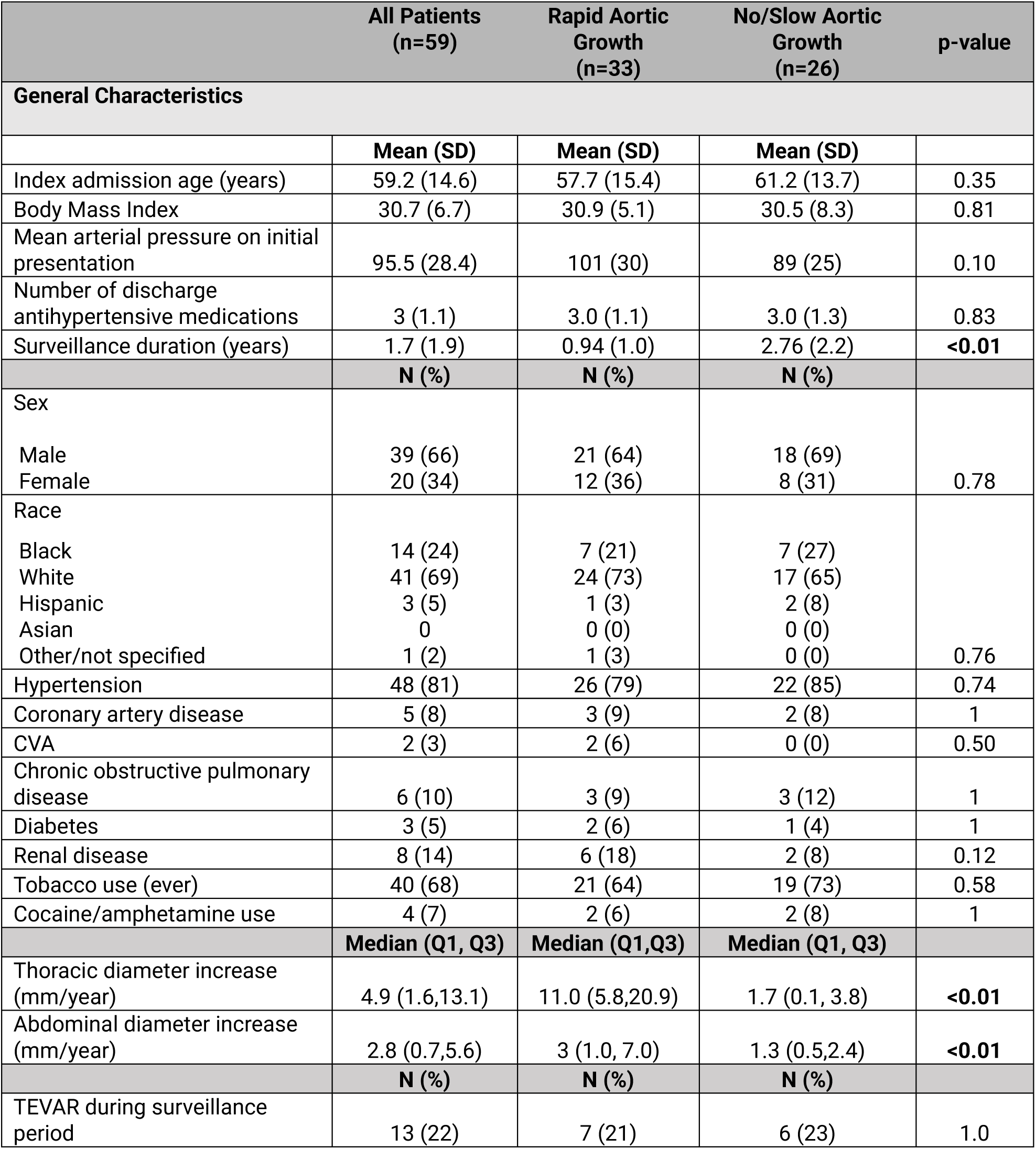
Patient demographics, comorbidities, aortic growth (mm/year), and TEVAR incidence during surveillance duration.

### Baseline CT characteristics

Baseline imaging characteristics are shown in **Table II**. Among all patients, baseline maximum thoracic aortic diameter was 42.0±7.0 mm and baseline maximum abdominal aortic diameter was 32.9±5.0 mm. Maximum thoracic aortic diameter was greater in patients with no/slow aortic growth <5mm/year (44.3±8.5 mm) compared to those with rapid aortic growth (40.3±5.0 mm) (p=0.03). Abdominal aortic diameter was also greater in patients with no/slow aortic growth (34.3±5.7) than those with rapid aortic growth (31.7± 4.1) (p=0.05). There were no differences in false lumen patency, incidence of false lumen diameter >22mm, or incidence of lesser curve entry tear between groups (p>0.05).

**Table II:** Imaging characteristics from initial CTA obtained at index admission.

|  | All Patients<br>(n=59) | Rapid Aortic<br>Growth<br>(n=33) | No/Slow Aortic<br>Growth<br>(n=26) | p-value |
| --- | --- | --- | --- | --- |
| **Baseline CT Characteristics** |  |  |  |  |
|  | **Mean (SD)** | **Mean (SD)** | **Mean (SD)** |  |
| Maximum Thoracic Aortic Diameter (mm) | 42.0 (7.0) | 40.3 (5.0) | 44.3 (8.5) | **0.03** |
| Maximum Abdominal Aortic Diameter (mm) | 32.9 (5.0) | 31.7 (4.1) | 34.3 (5.7) | **0.05** |
|  | **N (%)** | **N (%)** | **N (%)** |  |
| False Lumen Status |  |  |  |  |
| Thrombosed | 5 (8.5) | 1 (3.0) | 4 (15) |  |
| Partially Thrombosed | 27 (46) | 18 (55) | 9 (35) |  |
| Patent | 27 (46) | 14 (42) | 13 (50) | 0.13 |
| Lesser curve entry tear | 3 (5) | 2 (6) | 1 (4) | 1.0 |
| **Surveillance CT Characteristics** |  |  |  |  |
|  | **Mean (SD)** | **Mean (SD)** | **Mean (SD)** |  |
| Maximum Thoracic Aortic Diameter (mm) | 48.8 (9.2) | 48.8 (8.8) | 48.8 (10.0) | 1.0 |
| Maximum Abdominal Aortic Diameter (mm) | 38.7 (7.6) | 36.7 (6.7) | 38.7 (7.6) | 0.27 |
|  | **N (%)** | **N (%)** | **N (%)** |  |
| False Lumen |  |  |  |  |
| Thrombosed | 2 | 0 | 2 |  |
| Partially Thrombosed | 28 | 16 | 12 |  |
| Patent | 29 | 17 | 12 | 0.27 |

### Interval CT characteristics

Interval imaging characteristics are shown in **Table II**. At the interval CT scan, mean thoracic aortic diameter was similar in the no/slow growth group (48.8±8.8) compared to the rapid growth group (48.8±10.0) (p=1.0). There was no difference in abdominal aortic diameter between the no/slow growth group (36.7±6.7) and the rapid growth group (38.7±7.6) (p=0.27). There remained no differences in false lumen patency or false lumen diameter >22mm between groups (p>0.05).

### Volumetric Growth

**Table III** shows percentage of aortic volume increase by aortic zone over time. All measured zones of the aorta increased in volume over time, with the largest relative increase in Zone 5 with a median 24% increase (IQR 4.4-37) during the surveillance duration. Representative model output is shown in **Figure 4**. Compared to patients with no/slow growth, patients with rapid growth had higher percentage growth in all zones with significant differences in zone 4 (26% IQR 11-106 vs 4.1% IQR -2.2-13, p<0.01), zone 5 (32% IQR18-60 vs 5.8% IQR 0.69-24, p<0.01), and zone 9 (15% IQR -0.14-45 vs 0.56% IQR -9.6-4.6, p=0.01). **Figure 5** shows percentage growth over time by patient. While most patients experienced an increase in zone volume over time, there was significant variation by patient and by aortic zone.

**Figure 4:**
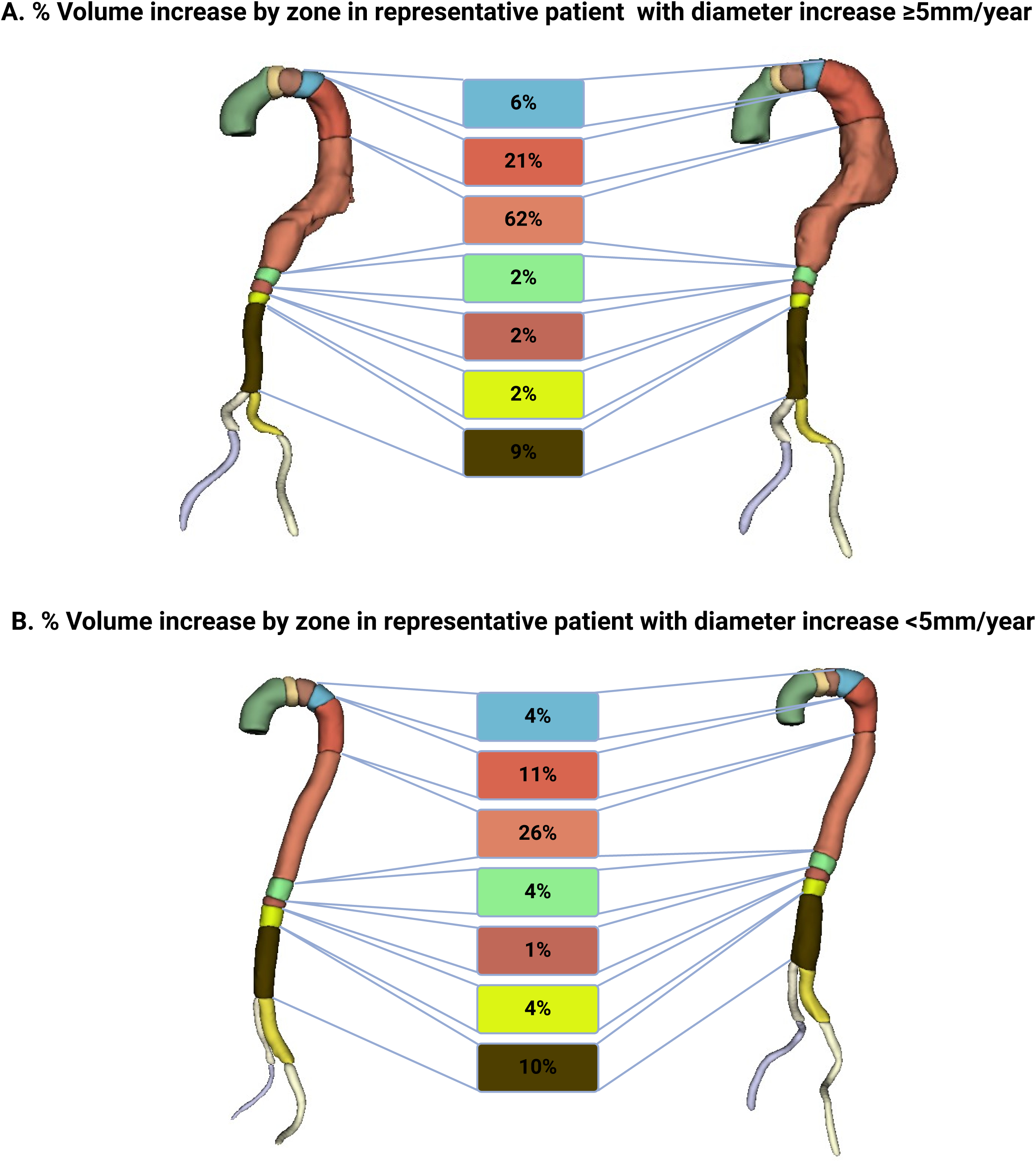
Changes in aortic zone volumes 3-10 between baseline (left) and surveillance (right) imaging in two representative patients with variable growth patterns. The deep learning model captured volumetric changes in aortic zones over time with zones 4 and 5 experiencing the largest relative increases in volume.

**Figure 5:**
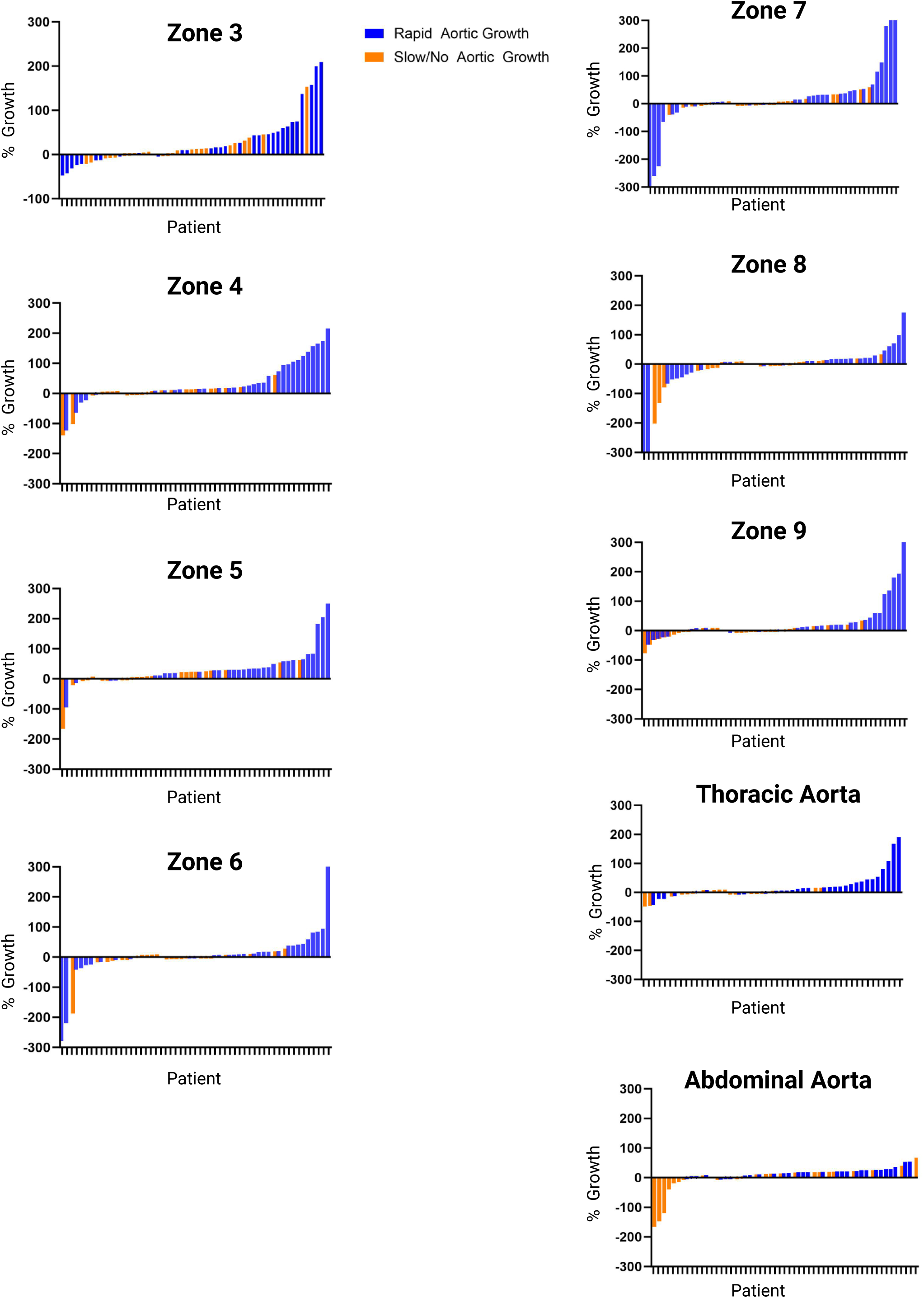
Waterfall plots demonstrating changes in percent volume increase over time by patient. Each bar corresponds to an individual patient, with y-axis representing percentage growth over time. Zones 3,4, and 5 had the largest relative increases in zone volume, while zones 6,7, and 8 were more variable. Patients in blue experienced rapid growth, while those in orange had no/slow growth.

**Table III:** Aortic growth over time by group defined as % dilation ((surveillance aortic volume – initial aortic volume)/initial aortic volume)*100).

|  | All Patients<br>(n=59) | Rapid Aortic<br>Growth<br>(n=33) | No/Slow Aortic<br>Growth<br>(n=26) | p-value |
| --- | --- | --- | --- | --- |
| **Aortic Growth Over Surveillance Duration (% dilation)** |  |  |  |  |
|  | **Median (Q1, Q3)** | **Median (Q1,Q3)** | **Median (Q1,Q3)** |  |
| Zone 3 Growth | 11 (-3.5,43) | 16 (-5.0,60) | 3.6 (-3.4,19) | 0.11 |
| Zone 4 Growth | 13 (2.0,35) | 26 (11,106) | 4.1 (-2.2,13) | <0.01 |
| Zone 5 Growth | 24 (4.4,37) | 32 (18,60) | 5.8 (0.69,24) | <0.01 |
| Zone 6 Growth | 4.0 (-9.9,17) | 9.5 (-10,39) | 0.57 (-8.5,4.0) | 0.42 |
| Zone 7 Growth | 4.3 (-4.3,34) | 27 (-9.5,49) | 2.8 (-3.6,9.7) | 0.55 |
| Zone 8 Growth | 2.6 (-17,17) | 10 (-29,21) | 0.93 (-14,5.8) | 0.40 |
| Zone 9 Growth | 4.5(-0.1,21) | 15 (-0.14,45) | 0.56 (-9.6,4.6) | 0.01 |
| Thoracic Aorta | 15 (1.2,22) | 18 (4.2,26) | 11 (-14, 19) | 0.02 |
| Abdominal Aorta | 5.3 (-1.1,19) | 15 (2.3,37) | -0.34 (-7.0,3.3) | 0.01 |

### Baseline Zone Volume Comparison

Mean aortic zone volumes obtained from baseline CTA are shown in **Table IV**. Mean zone 3 volume was greater in patients with no/slow aortic growth (6.0 voxels^3^ SD 1.4), compared to patients with rapid growth (5.0 voxels^3^ SD 1.8) (p=0.03). There were no other differences in baseline zone volumes between groups (p>0.05 for all).

**Table IV:** Baseline aortic zone volumes in voxels^3^ by group.

|  | All Patients<br>(n=59) | Rapid Aortic<br>Growth<br>(n=33) | No/Slow Aortic<br>Growth<br>(n=26) | p-value |
| --- | --- | --- | --- | --- |
| **Baseline Aortic Zone Volumes (voxels<sup>3</sup>)** |  |  |  |  |
|  | **Mean (SD)** | **Mean (SD)** | **Mean (SD)** |  |
| Zone 3 | 5.4 (1.7) | 5.0 (1.8) | 6.0 (1.4) | 0.03 |
| Zone 4 | 20.3 (7.4) | 19.4 (7.0) | 21.4 (7.9) | 0.32 |
| Zone 5 | 38.8 (17.1) | 37.0 (12.9) | 41.2 (21.5) | 0.38 |
| Zone 6 | 3.1 (1.7) | 2.8 (0.9) | 3.6 (2.3) | 0.07 |
| Zone 7 | 1.8 (0.9) | 1.7 (1.1) | 1.8 (0.8) | 0.84 |
| Zone 8 | 2.5 (1.5) | 2.2 (1.1) | 2.8 (2.0) | 0.19 |
| Zone 9 | 11.0 (3.9) | 10.5 (3.7) | 11.6 (4.0) | 0.28 |
| Thoracic Aorta | 64.5 (26.4) | 57.7 (23.8) | 65.8 (29.1) | 0.25 |
| Abdominal Aorta | 18.3 (8.1) | 16.1 (7.0) | 19.0 (7.8) | 0.15 |

### Segmentation model

The performance of the 3D-model is measured based on the number of overlapping pixels between the physician-annotated ground-truth (actual) region and the model predicted region. Performance is scored based on the Dice coefficient which ranges from 0 to 1 with 1 indicating a perfect match. Dice coefficient was tested using random sample of the training dataset with an overall performance of 0.73. Performance was best in Zone 4 (0.82), Zone 5 (0.88), and Zone 9 (0.91), with worse performance in Zone 3 (0.54), Zone 6 (0.60), Zone 7 (0.65), and Zone 8 (0.70). (**Table V)**

**Table V:** Dice coefficient by zone.

| Dice Coefficient by Zone |  |
| --- | --- |
|  | Dice Coefficient |
| Zone 3 | 0.54 |
| Zone 4 | 0.82 |
| Zone 5 | 0.88 |
| Zone 6 | 0.60 |
| Zone 7 | 0.65 |
| Zone 8 | 0.70 |
| Zone 9 | 0.91 |
| Overall | 0.73 |

## Discussion

To our knowledge, this is the first description of an automatic deep learning aortic segmentation model incorporating society-defined aortic zones and trained using a real-world dataset of patients with acute uncomplicated TBAD. The open-source, trained model demonstrates excellent concordance to a manually segmented “ground truth” aorta with the strongest performance in zones 4, 5, and 9. The model was able to accurately track changes in aortic volume over time and by patient, although in contrast to our hypothesis there were no differences in baseline zone volumes between the rapid aortic growth and no/slow aortic growth groups. This model provides a framework to accurately follow aortic volumes over time, with long-term potential for applications in growth prediction models, diagnosis and treatment planning, and incorporation into personalized treatment algorithms for dissection patients.

In vascular surgery, 3D image reconstruction has long played a role in preoperative planning and device selection. Contemporary commercially available software designed to assist with operative planning offers accurate reconstruction but is costly, semi-automatic, and requires extensive training thereby limiting its utility.^21^ For these reasons, machine learning, a subfield of artificial intelligence involving the development of algorithms capable of learning from data without programmed instruction, has generated interest for use in aortic pathology. In the context of aortic dissection, several studies have applied machine learning techniques to 3D image analysis, largely for the purpose of clinical diagnosis. Both Feiger et al. and Harris et al. developed machine learning models for the segmentation and classification of aortic dissection.^22–24^ Harris et al. demonstrated a sensitivity and specificity >94% for the diagnosis of aortic dissection based on a large sample of imaging studies obtained in a teleradiology practice, while the proposed model developed by Feiger et al. focused specifically on TBAD imaging and demonstrated high segmentation accuracy for identification of true lumen, false lumen, and total aorta.^22,23^ Other groups have proposed models for the diagnosis of abdominal aortic aneurysm (AAA) with diagnostic accuracy of up to 99%.^25–27^ These models collectively highlight the potential for machine learning applications in vascular surgery.

A similarity of these prior studies is that each model used CNN-based architecture. While a CNN-based approach is well suited for the task of solid organ segmentation or performing a binary operation like identifying the presence or absence of pathology, a CNN-based approach failed in our case when applied to the more complex task of aortic zone segmentation. In our dataset, initial attempts to perform aortic segmentation using existing CNN-based models resulted in object misidentification and over/under segmentation of multiple zones in nearly every patient.^20^ The CIS-UNet model we developed for this study builds off of the CNN-based approach by incorporating a novel self-attention block and vision transformer, both of which serve to improve segmentation accuracy in the task of aortic zone segmentation. Still, the overall dice coefficient of 0.73 across all zones leaves room for improvement. It is worse than scores attained with other models applied to simpler segmentation tasks: using CNN-based architecture, Yu et el found mean dice coefficient scores exceeding 0.93 for aorta, true lumen, and false lumen segmentation. ^24^ This highlights the added difficulty of incorporating aortic zones into a segmentation model and the opportunity for further refinement.

A principal application of this model is that it offers the opportunity to accurately follow aortic pathology in TBAD patients using automated zonal volumetric and diameter measurements.

Since the publication of updated TBAD reporting standards, only one study has analyzed zone-based growth of medically-managed TBAD. Blakeslee-Carter et al analyzed center-line diameter changes in standardized locations within each aortic zone over time in a similar cohort of 76 patients with medically managed acute/subacute TBAD.^28^ In this study, diameter growth was most pronounced in zones 3 (4.8±4.2 mm/year), 4 (4.7±4.4 mm/year), and 5 (3.7±3.5 mm/year), in agreement with our volume-based assessment, where growth was most pronounced in zones 3-5.^28^ This study showcases the potential application for a machine learning model like the one developed here to automate and expedite the task of taking manual measurements at specific locations within each aortic zone over time. Furthermore, the ability to generate both maximum diameter and volumetric data may allow for more robust growth analyses over time.

Another application of the proposed model is for the automation of TBAD classification. Previous models developed for aortic dissection or aneurysm have successfully performed the binary task of identifying the presence or absence of pathology, but have been unable to perform the more difficult task of differentiating between dissections based on a classification system, such as the Stanford, Debakey, or SVS/STS reporting standard systems.^9,23,29,30^ Rapid, automated determination of the dissection type could facilitate an accurate diagnosis and timely surgical consultation to the appropriate team, leading to life saving treatment. While efforts are ongoing to incorporate segmentation of the true and false lumens of the dissection into our current model, when added to the existing zone-based framework the model could provide a 3D assessment of the dissection flap morphology along with its proper classification according to the extent of the dissection and the affected zones.

The ultimate goal is to incorporate the data generated from the model output into a growth prediction algorithm that can assist with the selection of the most appropriate management pathway for patients with acute TBAD. In AAA, machine learning models have outperformed logistic regression in models predicting disease prevalence and mortality after rupture, and been used to predict annual aortic growth to within 2mm at 85% accuracy.^31–34^ Compared to AAA, TBAD may be an inherently more complex pathology than AAA to implement into a growth prediction algorithm due to variable patterns of dissection morphology and disease extent, and current understanding of high-risk radiographic parameters is limited. Conventional radiographic criteria defined as high risk in the literature, including aortic diameter >40mm, false lumen diameter >22mm, and lesser cure entry tear are based on small retrospective analyses and have not been prospectively validated.^9^ The automated model we propose offers the capability to readily attain these discrete measurements at a granular level. While our initial hypothesis that baseline zone volumes would be associated with aortic growth over time was unfounded, we were limited by the small number of patients available for analysis. It is worth noting that when looking at other conventional high-risk features, there were also no differences between groups, which speaks to the limitations of our sample size and the need for further testing and model optimization.

### Limitations

There are several other important limitations to consider. We excluded patients with aberrant anatomy to optimize model performance but further efforts will need to incorporate anatomic variation. Model performance was suboptimal in visceral segment zones where significant variations exist with celiac, superior mesenteric, and renal artery takeoffs. With exposure to additional imaging and anatomic variation of the visceral and renal vessels, the performance in zones 6 through 8 is expected to improve. The sample also had a considerable number of patients with an only three-month interval between baseline and surveillance CT. This limited our ability to analyze long-term growth in many patients, and as a result, our sample may have been biased towards patients that experienced early rapid aortic growth. Additionally, there is not a standard definition of aortic growth in the literature, which limits our ability to compare growth patterns and characteristics between work from other similar studies. Our model also did not consider dissection flap morphology or false lumen volume. The next model iteration will add these components to the existing zone segmentation to better understand their contributory role, if any, towards aortic growth over time and allow for a more comprehensive set of applications with the existing model framework.

## Conclusions

This is the first description of an automatic deep learning aortic segmentation model incorporating SVS-defined aortic zones and trained using a real-world dataset of patients with uncomplicated Type B aortic dissection. The open-source, trained model demonstrates high concordance to manually segmented “ground truth” aorta with particularly strong performance in zones 4, 5, and 9. Volumetric enlargement was most pronounced in Zones 4 and 5. The model framework offers potential to rapidly obtain volumetric and diameter measurements that may allow for a robust and granular understanding of aortic behavior. In our limited sample, there did not appear to be differences in baseline aortic zone volumes between patients with and without aortic enlargement.

## Data Availability

All data produced in the present study are available upon reasonable request to the authors

## Author Contributions

Conception and design: JK, BF, WS, MC. Data collection: JK, BF, CV, BB, GS. Statistical analysis and interpretation: JK, BF, MC, MI. Writing the article: JK, MI. Critical revisions: BF, MC, GRU. Final approval: JK, MI, BF, CV, BB, GS, EH, GRU, WS, MC

